# UV-C irradiation is highly effective in inactivating SARS-CoV-2 replication

**DOI:** 10.1101/2020.06.05.20123463

**Authors:** Mara Biasin, Andrea Bianco, Giovanni Pareschi, Adalberto Cavalleri, Claudia Cavatorta, Claudio Fenizia, Paola Galli, Luigi Lessio, Manuela Lualdi, Enrico Tombetti, Alessandro Ambrosi, Edoardo Maria Alberto Redaelli, Irma Saulle, Daria Trabattoni, Alessio Zanutta, Mario Clerici

**Affiliations:** Department of Biomedical and Clinical Sciences L. Sacco, University of Milano, Milano, Italy; Italian National Institute for Astrophysics (INAF) – Brera Astronomical Observatory, Merate, Italy; Epidemiology and Prevention Unit, IRCCS Foundation, Istituto Nazionale dei Tumori, Milan, Italy; Italian National Institute for Astrophysics (INAF) – Padova Astronomical Observatory, Padova, Italy; Department of Imaging Diagnostic and Radioterapy, IRCCS Foundation, Istituto Nazionale dei Tumori, Milan, Italy; University Life and health San Raffaele, Milan, Italy; Department of Pathophysiology and Transplantation, University of Milano, Milano, Italy; Don C. Gnocchi Foundation, IRCCS Foundation, Milano, Italy

## Abstract

The potential virucidal effects of UV-C irradiation on SARS-CoV-2 were experimentally evaluated for different illumination doses and virus concentrations (1000, 5, 0.05 MOI). At a virus density comparable to that observed in SARS-CoV-2 infection, an UV-C dose of just 3.7 mJ/cm^2^ was sufficient to achieve a more than 3-log inactivation without any sign of viral replication. Moreover, a complete inactivation at all viral concentrations was observed with 16.9 mJ/cm^2^. These results could explain the epidemiological trends of COVID-19 and are important for the development of novel sterilizing methods to contain SARS-CoV-2 infection.

## Introduction

The COVID-19 pandemic caused by SARS-CoV-2 virus^1^ has had an enormous, as yet barely understood, impact on health and economic outlook at the global level^2^. The identification of effective microbicide approaches is of paramount importance in order to limit further viral spread, as the virus can be transmitted via aerosol^3,4^ and can survive for hours outside the body^5–7^. Non-contact disinfection technologies are highly desirable, and UV radiation, in particular UV-C (200 – 280 nm) has been suggested to be able to inactivate different viruses, including SARS-CoV^8–12^. The interaction of UV-C radiations with viruses has been extensively studied^13–15^, and direct absorption of the UV-C photon by the nucleic acid basis and/or capsid proteins leading to the generation of photoproducts that inactivate the virus was suggested to be one of the main UV-C-associated virucidal mechanisms^16,17^. Some models have been proposed to correlate the nucleic acid structure with the required dose to inactivate the virus, but a reliable model is still unavailable^18^. This is also due to the fact that UV-C measurements were conducted using different viruses and diverse experimental conditions^19–22^. This led to an extremely wide range of values for the same virus and, e.g., in the case of SARS-CoV-1 values reported in the literature range from a few mJ/cm^2^ to hundreds mJ/cm^2 19,22,23^. Likewise, recent papers reported values for UVC inactivation ranging from 3 to 1000 mJ/cm^2 24–27^. A better understanding of the effects of UV-C on SARS-CoV-2, which take into account all the key factors involved in the experimental setting (including culture medium, SARS-CoV-2 concentration, UV-C irradiance, time of exposure, and UV-C absorbance) will allow to replicate the results in other laboratory with different devices. Moreover, as recent evidences suggest that UV light from sunlight are efficient in inactivating the virus^28^, these measurements will be relevant for the setting up of further experiment considering the role of UV-A and UV-B on SARS-CoV-2 replication.

## Results and discussion

Herein, we report the effect of monochromatic UV-C (254 nm) on SARS-CoV-2, showing that virus inactivation can be easily achieved. Experiments were conducted using a custom-designed low-pressure mercury lamp system, which has been spectral-calibrated providing an average intensity of 1.082 mW/cm^2^ over the illumination area (see the details reported in the Method section). Three different illumination exposure times, corresponding to 3.7, 16.9 and 84.4 mJ/cm^2^, were administered to SARS-CoV-2 either at a multiplicity of infection (MOI) of 0.05, 5, 1000. The first concentration is equivalent to the low-level contamination observed in closed environments (e.g. hospital rooms), the second one corresponds to the average concentration found in the sputum of COVID-19 infected patients, and the third one is a very large concentration, corresponding to that observed in terminally diseased COVID-19 patients^29^. After UV-C exposure, viral replication was assessed by culture-polymerase chain reaction (C-RT-PCR) targeting two regions (N1 and N2) of the SARS-CoV-2 nucleocapsid gene, as well as by analyzing SARS-CoV-S2-induced cytopathic effect. Analyses were performed in the culture supernatant of infected cells at three different time points (24, 48 and 72 hours for SARS-CoV-2 at MOI 1000 and 5; 24, 48 hours and 6 days for SARS-CoV-at MOI 0.05), as well as on cell lysates at the end of cellular culture (72 hours: MOI 1000 and 5; 6 days: MOI 0.05). This approach allows to follow the kinetic of viral growth and to verify whether the used dose is sufficient to completely inactivate the virus over time. This is useful from a practical point of view, when UV-C devices are used to disinfect surfaces and the environment.

The effect of the UV-C exposure on SARS-CoV-2 replication was extremely evident and independent from the MOI employed; dose-response and time-dependent curves were observed. Figure 1, 2 and 3 report for different MOI the number of SARS-CoV-2 copies for the three concentrations as a function of the UV-C dose and time, quantified on a standard curve from a plasmid control. The corresponding normalised curves of the virus copies are reported in the same figures.

**Figure 1.**
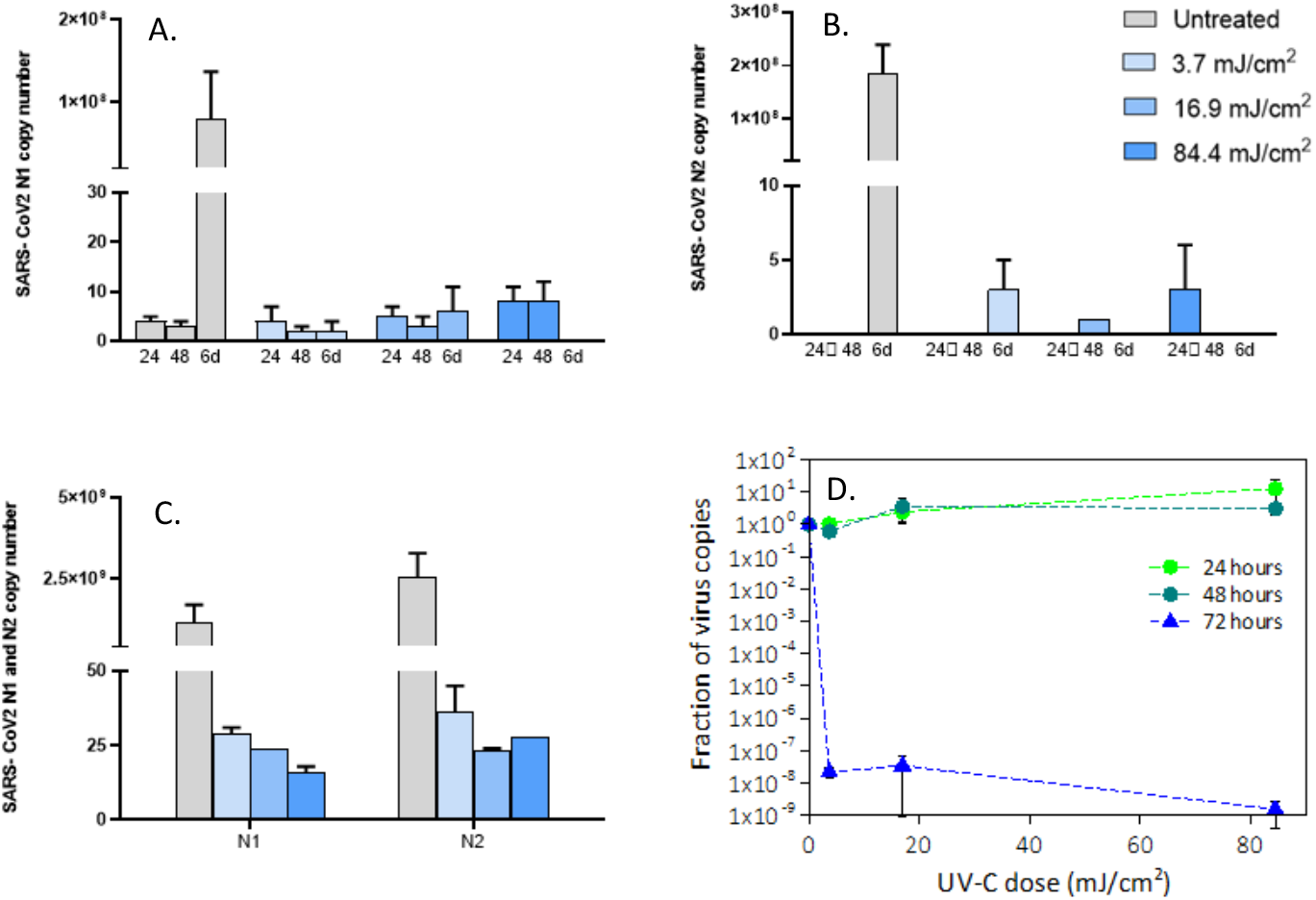
Viral replication of UV8irradiated SARS8CoV82 (0.05 MOI) virus in in vitro VeroE6 cells. Vero E6 cells were infected with UV-C irradiated SARS-CoV-2 virus at a MOI of 0.05. Culture supernatants were harvested at the indicated times (24, 48 hours and 6 days) and virus titers were measured (Panel A, B) by absolute copy number quantification (Real-Time PCR). Viral replication was assessed even on cell lysate harvested at the end of cell cultures (6 days) (Panel C). All cell culture conditions were seeded in duplicate. Panel D reports the plots of the measured virus copies normalized at the untreated sample in the different conditions. For descriptive purposes, mean values and whiskers representing the observed half-ranges are shown.

**Figure 2.**
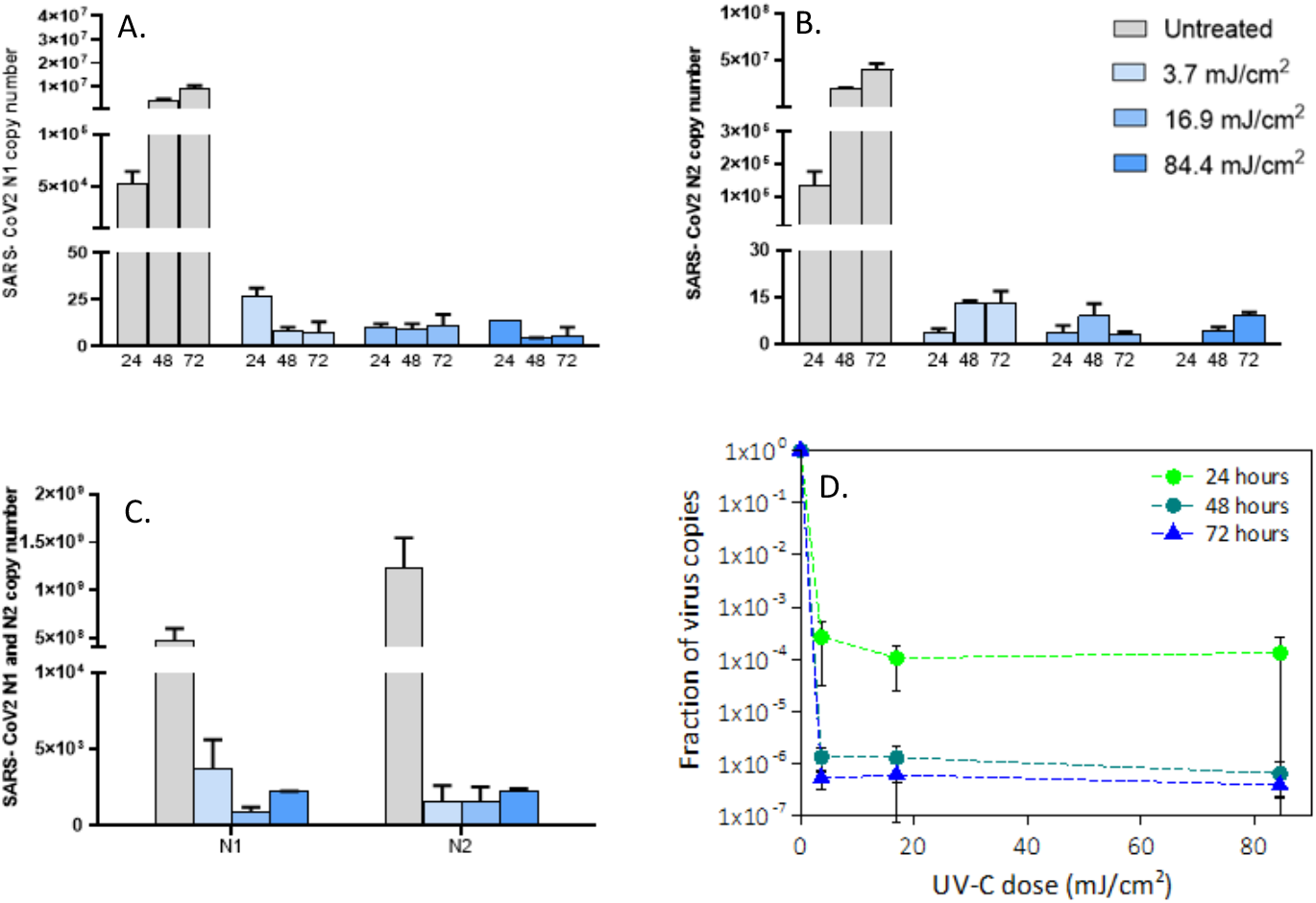
Viral replication of UV8irradiated SARS8CoV82 (5 MOI) virus in in vitro VeroE6 cells. Vero E6 cells were infected with UV-C irradiated SARS-CoV-2 virus at a MOI of 5. Culture supernatants were harvested at the indicated times (24, 48 and 72 hours) and virus titers were measured by absolute copy number quantification (Real-Time PCR, A and B). Viral replication was assessed even on cell lysate harvested at the end of cell cultures (72 hours) (C). All cell culture conditions were seeded in duplicate. Panel D reports the plots of the measured virus copies normalized at the untreated sample in the different conditions. For descriptive purposes, mean values and whiskers representing the observed half-ranges are shown.

**Figure 3.**
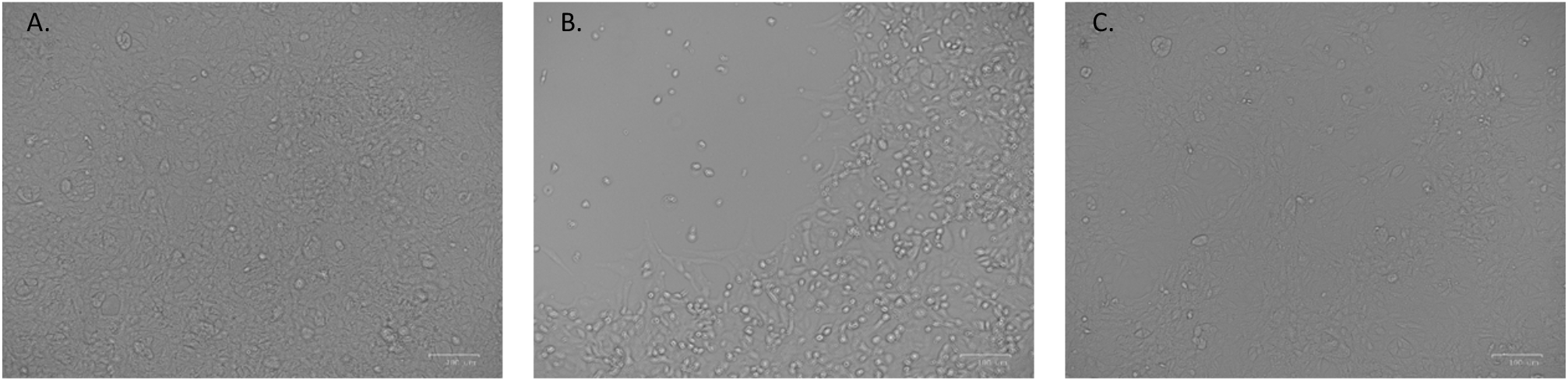
Analyses of virus induced cytopathic effect. (A) No cytopathic effect was observed in uninfected cultured VeroE6 monolayers maintained in 50mJ/cm^2^ UV-treated complete medium for 72 hours. (B) In vitro infection of SARS-CoV-2 (5 MOI) UV-C untreated VeroE6 cells resulted in an evident cytopathic effect. (C) SARS-CoV-2 irradiation with 3.7 mJ/cm^2^ UV-C rescued the cytopathic effect induced by UV-C untreated virus.

Viral replication was not observed at the lowest viral concentration (0.05 MOI) in either untreated or in UV-C-irradiated samples in the initial 48 hours (Figure 1). However, 6 days after infection, viral replication was distinctly evident in the UV-C unexposed condition, but was completely absent following UV-C irradiation even at 3.7 mJ/cm^2^ both in cell culture supernatants (Figure1, panel A and B) and in cell lysate (Figure 1, panel C). A two-way ANOVA analysing the effect of UV-C dose and time of incubation failed to identify a significant effect of the UV exposure on viral replication. This is due to the fact that at very low MOI relevant increases in N1 and N2 copy numbers were detectable only in a single condition -at six days in the absence of UV-C exposure-thus hampering the statistical power of the analysis.

At the intermediate viral concentration (5 MOI), a significant reduction of copy number starting from the 3.7 mJ/cm^2^ dose with a decrease of a factor of 2000 (> 3-log decrease) after 24 hours was observed (Figure 2, panel D). A two-way ANOVA confirmed that this UV-C dose significantly dampened viral replication (p=0.000796, and P=0.000713 for N1 and N2 copies respectively). Even more important, the copy number did not increase over time, suggesting an effective inactivation of the virus, which was further confirmed by cytopathic effect assessment (Figure 3, panel A, B and C).

Using a high viral input (MOI=1000), the two-way ANOVA confirmed that all the tree UV doses analysed resulted in a significant suppression of viral replication for both N1 (3.7 mJ/cm^2^: p= 0.008455; 16.9 mJ/cm^2^: p=0.004216; and 84.4 mJ/cm^2^: p=0.000202) and N2 copies (3.7 mJ/cm^2^: p= 6.43E-05; 16.9 mJ/cm^2^: p=1.68E-05; and 84.4 mJ/cm^2^: p=1.68E-05)(Figure 4). Notably, a different course of infection was observed, in which the inhibitory effect was not accompanied by viral suppression for the UV-C dose of 3.7 mJ/cm^2^ (Figure 4, panel A, B and D). Indeed, a relevant reduction in N1 e N2 copy numbers was observed in a UV-C dose-dependent manner as early as 24 hours (by a factor of 10^3^ at 3.7 mJ/cm^2^ and 10^4^ at 16.9 mJ/cm^2^, Figure 4, panel A, B and D), but longer culture times resulted in an increase in N1 and N2 copy numbers for the UV-C dose of 3.7 mJ/cm^2^. This indicates that the residual viral input left by the 3.7 mJ/cm^2^ was able to replicate and suffucient to generate an effective infection. This is not the case in cultures exposed to higher UV-C doses, as no replication could be detected in these conditions. All the results were further confirmed by 2-ANOVA statistical analyses performed on viral replication at intracellular level (3.7 mJ/cm^2^ vs. Untreated: N1, p=0.008455; N2: p=6.43E-05; 16.9 mJ/cm^2^ vs. Untreated: N1, p=0.004216; N2: p=1.68E-05; 84.4 mJ/cm^2^ vs. Untreated: N1, p=0.004216; N2: p=1.68E-05) (Figure 1, panel C; Figure 2, panel C; Figure 4, panel C).

**Figure 4.**
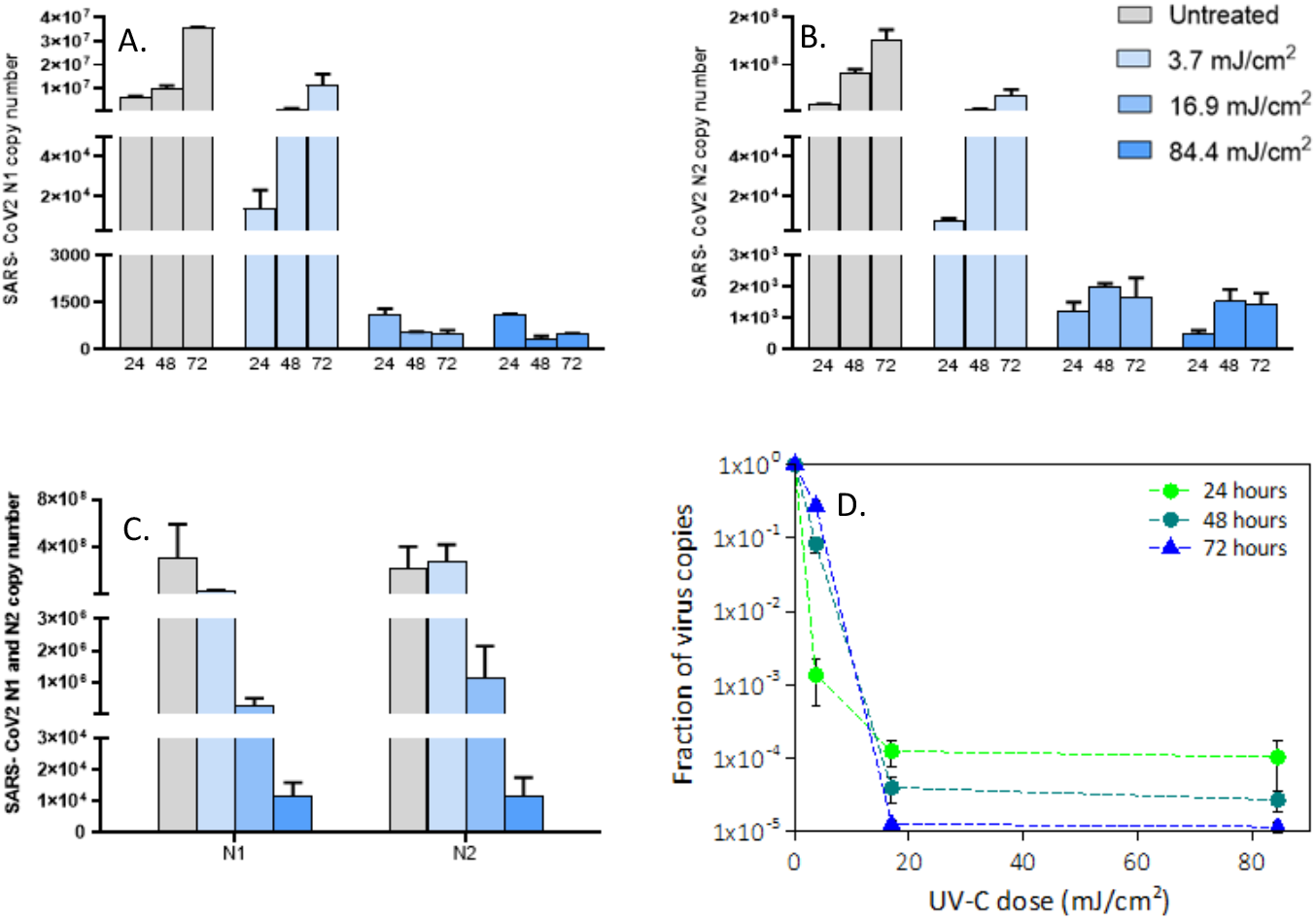
Viral replication of UV8irradiated SARS8CoV82 (1000 MOI) virus in in vitro VeroE6 cells. Vero E6 cells were infected with UV-C irradiated SARS-CoV-2 virus at a MOI of 1000. Culturesupernatants were harvested at the indicated times (24, 48 and 72 hours) and virus titers were measured (Panel A, B) by absolute copy number quantification (Real-Time PCR). Viral replication was assessed even on cell lysate harvested at the end of cell cultures (72 hours) (Panel C). All cell culture conditions were seeded in duplicate. Panel D reports the plots of the measured virus copies normalized at the untreated sample in the different conditions. For descriptive purposes, mean values and whiskers representing the observed half-ranges are shown.

We compared our results with data available in the literature and observed that our inactivating dose is much smaller than that reported in Heilingloh et al.^26^ (1000 mJ/cm^2^ for the complete inactivation). This discrepancy is likely to be the consequence of the UV-C absorption by the medium used in Heilingloh et al, which has a 4-fold higher thickness compared to the one used in our experiments. This possibility is supported by the observation that 200 mJ/cm^2^ of UV-A, which is not absorbed by the medium, was sufficient to reduce viral replication of 1-log. As UV-A light is significantly less efficient (order of magnitudes) than UV-C, the reported UV-C inactivating dose (100mJ/cm2) seems to be questionable. Two other papers measured the effect of UV-C light on SARS-CoV-2. In Ruetalo et al.^25^, the illumination of 254 nm light was employed on a dried sample of SARS-CoV-2. Complete inactivation was obtained with 20 mJ/cm^2^, a value greater than ours, but in the same range. It has to be underlined that in the dried film a shielding effect by the organic component present in the liquid can occur, reducing the efficiency of the UV-C light. This was shown by Ratnesar-Shumate et al.^28^, who demonstrated that the dose required to obtain a similar degree of viral inactivation was twice in dried samples from gMEM compared to the ones resuspended in simulated saliva. Notably, the two mediums differ for their composition, mainly in terms of protein and solid percentage, with higher values for the gMEM.

Inagaki et al. used a 285 nm UV LED and showed that a dose of about 38 mJ/cm^2^ was sufficient to completely inactivate SARS-CoV-2. This dose is greater compared to the one we established; this discrepancy can be explained by the observation that the 285 nm is less efficient than the 254 nm wavelength^24^. Finally, in an elegant study Storm et al.^27^ compared the virucidal effect of UV-C in wet and dry systems. Results were based on the use of a very small volume of viral stock in DMEM (5 μl) and showed that a dose of 3.4 mJ/cm^2^ inactivated wet samples, whereas a dose that twice as high was needed in dried samples. These results are comparable to the ones herein, and the shielding effect in dried samples is almost evident. Such comparisons show how the experimental conditions adopted significantly impact on the definition of the dose of UV-C resulting in virus inactivation. It is therefore crucial to accurately describe all the details of the experiments to perform a reliable comparison.

In conclusion, we report the results of a highly controlled experimental model that allowed us to identify the UV-C radiation dose sufficient to inactivate SARS-CoV-2. The response depends on both the UV-C dose and the virus concentration. Indeed, for virus concentrations typical of low-level contaminated closed environment and sputum of COVID-19 infected patients, a very small dose of less than 4 mJ/cm^2^ was enough to achieve full inactivation of the virus. Even at the highest viral input concentration (1000 MOI), viral replication was totally inactivated with a dose ≥16.9 mJ/cm^2.^ These results show how the SARS-CoV-2 is extremely sensitive to UV-C light and they are important to allow the proper design and development of efficient UV based disinfection methods to contain SARS-CoV-2 infection.

## Methods

### In vitro SARS8CoV82 infection assay

3 × 10^5^ VeroE6 cells were cultured in DMEM (ECB7501L, Euroclone, Milan, Italy) with 2 % FBS medium, with 100 U/ml penicillin and 100 μg/ml streptomycin, in a 24-well plate one day before viral infection assay. SARS-CoV-2 (Virus Human 2019-nCoV strain 2019-nCoV/Italy-INMI1, Rome, Italy) at a multiplicity of infection (MOI) of 1000, 5 and 0.05 were treated with different doses of UV-C radiation (see the dedicated section) before inoculum into VeroE6 cells. UV-C-untreated virus served as positive controls. Cell cultures were incubated with the virus inoculum in duplicate for three hours at 37°C and 5% CO_2_. Then, cells were rinsed three times with warm PBS, replenished with the appropriate growth medium and observed daily for cytopathic effect. Viral replication in culture supernatants was assessed by an Integrated Culture-polymerase chain reaction (C-RT-PCR) method^30^ at 24, 48, and 72 hours post-infection (hpi) while infected cells were harvested for RNA collection at 72 hpi. Cell cultures from SARS-CoV-2 at 0.05 MOI were harvested 6 days post infection. RNA was extracted from VeroE6 cell culture supernatant and cell lysate by the Maxwell® RSC Instrument with Maxwell® RSC Viral Total Nucleic Acid Purification Kit (Promega, Fitchburg, WI, USA), quantified by the Nanodrop 2000 Instrument (Thermo Scientific) and purified from genomic DNA with RNase-free DNase (RQ1 DNase; Promega). One microgram of RNA was reverse transcribed into first-strand cDNA in a 20-μl final volume as previously described^31,32^.

Real-time PCR was performed on a CFX96 (Bio-Rad, CA, USA) using the 2019-nCoV CDC qPCR Probe Assay emergency kit (IDT, Iowa, USA), which targets two regions (N1 and N2) of the nucleocapsid gene of SARS-CoV-2. Reactions were performed according to the following thermal profile: initial denaturation (95°C, 10 min) followed by 45 cycles of 15 s at 95°C (denaturation) and 1 min at 60°C (annealing-extension). Viral copy quantification was assessed by creating a standard curve from the quantified 2019-nCoV_N positive Plasmid Control (IDT, Iowa, USA).

### UV illumination test

The illumination of the virus solution was conducted using a low-pressure mercury lamp mounted in a custom designed holder, which consist in a box with a circular aperture 50 mm in diameter placed at approximately 220 mm from the source. The aperture works as a spatial filter to make the illumination of the area behind more uniform. A mechanical shutter is also present to start the illumination process. The plate is placed 30 mm below the circular aperture and a single dwell (34.7 mm in diameter), centered in respect to the 50 mm aperture, has been irradiated from the top. The dwell was filled with 0.976 ml of the virus suspended in Dulbecco’s Modified Eagle’s Medium (DMEM) in order to have a 1 mm thick liquid layer. After the irradiation, the sample was treated as described in the previous section.

The intensity of the lamp and its spectral properties have been measured using an Ocean Optics HR2000+ spectrometer (Ocean Optics Inc., Dunedin, USA). The HR2000+ spectrometer was calibrated against a reference deuterium–halogen source (Ocean Optics Inc. Winter Park, Winter Park, Florida) and in compliance with National Institute of Standards and Technology (NIST) practices recommended in NIST Handbook 150-2E, Technical guide for Optical Radiation Measurements. The last calibration was performed in March 2019. The detector of our spectrometer is a high-sensitivity 2048-element Charge-Coupled Device (CCD) array from Sony. The spectral range is 200–1100 nm with a 25 μm wide entrance slit and an optical resolution of 1.4 nm (FWHM). The cosine-corrected irradiance probe, model CC-3-UV-T, is attached to the tip of a 1 m long optical fibre and couples to the spectrometer. The intensity of the lamp has been measured by positioning the spectrometer in five positions: in the center and at the ends of a 20 mm cross arm after a warming up time of 30 s. The spectra in the five positions are reported in Figure 5 (Panel A) together with a scheme of the dwell and illuminated area.

**Figure 5.**
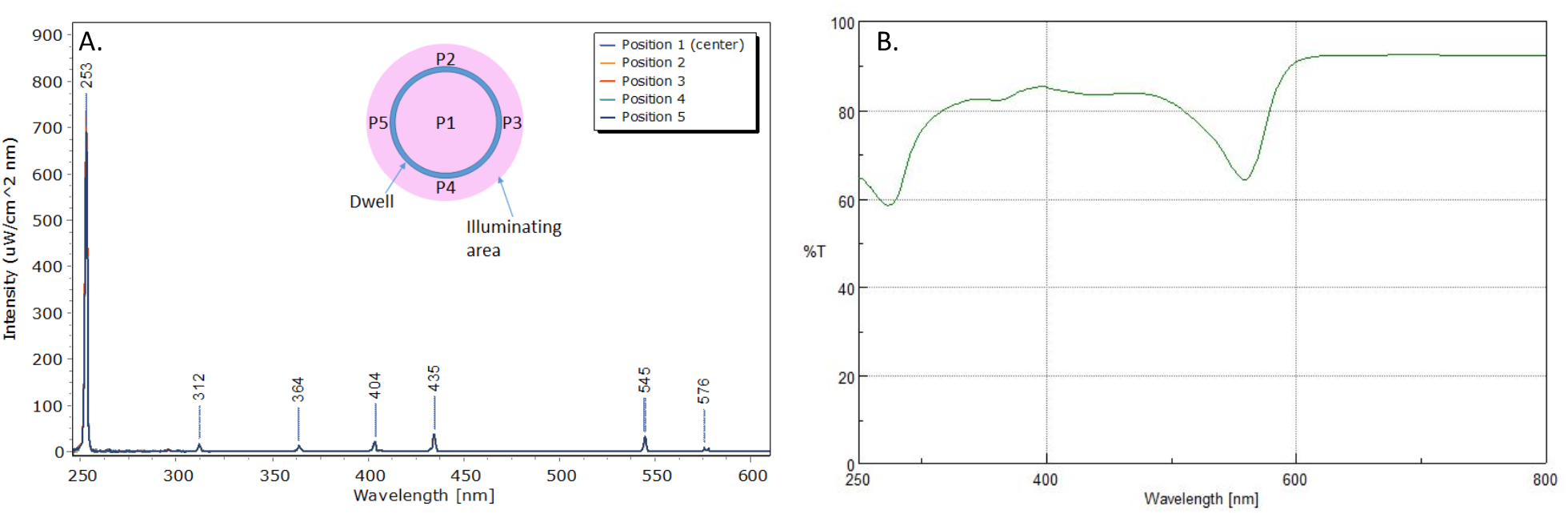
Mercury lamp spectrum measured in the five positions (Panel A). Inset: scheme of the illuminated dwell and the measuring position. UVXvis transmission spectrum of the Dulbecco’s Modified Eagle’s Medium (DMEM) in a 1 mm quartz cuvette (Panel B).

As expected, the emission is dominated by the UV-C line (Figure 5, Panel A) and its intensity was uniform in the area with an average value of 1.082 mW/cm^2^. The stability of the lamp was evaluated in +/-11E-3 mW/cm^2^ during a 130 s measurement. According to this value, three exposure times were set: 5, 23 and 114 s (with an accuracy of 0.2s), which correspond to following doses: 5.4, 25.0, 123.4 mJ/cm^2^. This is the nominal UV doses provided to the dwell, but we were interested in the effective doses (D_e_) reaching the virus. It was necessary to calculate the effective irradiance (I_e_). This step was performed considering both the reflection losses at the air/water interface (*R*_*w*_) and the Transmittance (*T*_*s*_) of the DMEM solution at 254 nm (from the spectrum in figure 5, Panel B, considering the cuvette losses, T_s_ = 0 .70). It is important to notice that the spectrum was measured in a quartz cuvette (1 mm thick) by means of a Jasco V770 spectrophotometer and this thickness was the same of the solution in the dwell during the UV irradiation step.

The reflection loss was computed as follow:

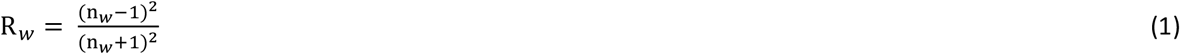

Where n_w_ = 1.375 is the refractive index of water at 254 nm. Then, *I*_*e*_ was calculated:

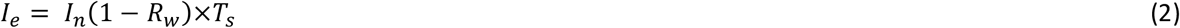

The final transmission of the DMEM solution was equal to 0.68 and the corresponding effective doses were derived simply multiplying *I*_*e*_ by the exposure time.

According to this value, the effective doses provided to the viruses were: 3.7±0.15, 16.9±0.2 and 84.4±0.9 mJ/cm^2^. We have to notice that we are neglecting here the absorption of the virus at this wavelength and the possible scattering. Such approximations are valid considering the relative low concentration of the virus and small thickness of the layer (the solution appeared fully transparent).

### Statistical analyses

To assess the effect of the different UV-C doses on N1 and N2 copy numbers, two-way ANOVAs were performed. For the analysis of intracellular N1 and N2 doses in the supernatant, UV-C dose and MOI represented the dependent variables, while for the analysis of N1 and N2 in the supernatant, different analyses were performed for individual MOI, using UV-C dose and time as dependent variables.

## Data Availability

The results are reported in the paper and in the annexed supplementary material

## Acknowledgements

This research was partially supported by a grant from Falk Renewables and it has been carried out in the context of the activities promoted by the Italian Government and in particular, by the Ministries of Health and of University and Research, against the COVID-19 pandemic. Authors are grateful to INAF’s President, Prof. N. D’Amico, for the support and for a critical reading of the manuscript.

## Author Contribution

P.G., L.L. E.R. and A.Z. designed and produced the illumination system; A.C., C.C. and M.L. performed the lamp calibration; A.B. performed the lamp setup and lamp dosimetry, wrote the main manuscript; M.B. performed biological experiments, analyzed the data, wrote the main manuscript; C.F., I.S. designed and performed some biological tests; E.T, A.A. performed the statistical analysis; D.T. discussed the results; G.P. and M.C. supervised the study and review the manuscript.

